# The relationship between the prevalence and severity of Non-suicidal self-injury (NSSI) and personality traits among Chinese junior high school students

**DOI:** 10.1101/2022.05.26.22275643

**Authors:** Zhongliang Jiang, Zhiyi Wang, Miao Zhao, Long He, Qiang He, Xiaojing Cheng, Jie Chen, Geng Tian, Xianbin Li, Muhammad Imran Haider, Jintong Liu

## Abstract

**Background:** Non-suicidal self-injury (NSSI) is more common in adolescents and its occurrence may be influenced by personality traits. Children and adolescents are a high-risk group for NSSI. Although there are some studies on the prevalence of NSSI, however, there are fewer studies on the factors associated with the severity of NSSI and the specific causes of the influence of NSSI and personality traits.

**Methods:** Participants of this study were junior high school students enrolled in three different schools of a Chinese province. NSSI was evaluated using the Adolescents Self-Harm Scale and their personality traits were assessed using the Neuroticism Extraversion Openness Five-factor Inventory (NEO-FFI).

**Results:** 2376 junior high school students participated, and the annual incidence of NSSI was 37.1% (n=881). The mean age of the NSSI-detected individuals was 13.61 years (SD=1.017). Out of total number of detections, 56.6% (n=499) were female and 67.4% (n=594) were individuals who self-injured using multiple means. Hair pulling, scratching the skin, and whacking harder objects with the hand were the most common modes of NSSI, with incidences of 51.0%, 43.0%, and 42.8% respectively in the NSSI-detected population. There was a negative association between grade and NSSI severity (p < 0.05), and NSSI was more severe in females. The scores in the Neuroticism dimension were higher in the group with NSSI than in the group without NSSI, while the scores in the extraversion, openness, agreeableness and conscientiousness dimensions were lower than in the group without NSSI (p < 0.01). Neuroticism and openness were significantly positively correlated with NSSI severity, and extraversion, agreeableness, conscientiousness were significantly negatively correlated with NSSI severity (p < 0.01).

**Conclusion:** The severity of NSSI was relatively higher in lower grades among junior grades and females, and the number of self-injury using multiple means was significantly higher; hair pulling, skin scratching and whacking harder objects with hands were the most common NSSI modalities. Among the personality traits, for the incidence of NSSI, high neuroticism was a risk factor, and high extraversion, openness, agreeableness, and conscientiousness were protective factors. For the severity of NSSI, high neuroticism and high openness were risk factors, and high extraversion, high agreeableness, and high conscientiousness were protective factors.

## Introduction

Non-suicidal self-injury (NSSI) is generally defined as intentional injury to one’s body, directly or indirectly, without explicit suicidal intent [1], which is neither intended to cause death nor socially acceptable [2]. The Diagnostic and Statistical Manual of Mental Disorders Fifth edition (DSM-5) states that NSSI most commonly begins in the early 10 years of age and persists for many years, and is essentially characterized by individuals repeatedly causing superficial but painful body surface injuries, most often with the aim of reducing negative emotions. On average, individuals with NSSI have been reported to self-injure in 6.2 ± 2.67 ways [3], with common ones including cutting, scratching, and hair pulling.

NSSI is repetitive, with 55.1% of individuals with NSSI continuing to have in the following 6 months [4]. Also, the individuals with NSSI have a higher risk of suicide [5], and NSSI often precedes suicide [6]. It is estimated that suicide rates are hundreds of times higher in individuals who self-injure than in the normal population [7]. However, NSSI is fundamentally different from suicide in terms of intent and lethality, etc. Studies have shown [8,9] that individuals who experience NSSI cognitively distinguish between NSSI and suicidal behavior, and they use NSSI as a coping strategy for the purpose of regulating negative emotions, among others. The experience avoidance model proposed by Chapman AL et al [10] suggests that a series of negative emotions induced by external stimuli, when individuals are unable to perform effective expression or have difficulty in emotional control, they resort to self-injurious behaviors to avoid or alleviate unpleasant and disgusting experiences, and this approach reinforces the relationship between NSSI and negative emotions in a chain reaction. In addition, the stress exposure model in psychopathology [11] similarly proposes a mechanism for the role of NSSI as a method to alleviate negative emotions. Children and adolescents are at high risk for NSSI, and numerous studies have shown that they are a high-risk group for NSSI [12]. Approximately 20% of individuals report onset before the age of 25 years [13]. According to incomplete statistics [14], the detection rate of NSSI among adolescents ranges from 13% to 46.5% in the United States, 17% in Canada, and 6.2% in Australia. The detection rate among adolescents in European countries ranged from 17.1% to 38.6% [15], and among Chinese secondary school students, the detection rate ranged from 5.4% [16] to 44.3% [17]. In recent years, NSSI has gradually become one of the research hotspots, and although there is some research on the incidence of NSSI, there is less research on the factors associated with the severity of NSSI, especially in the Chinese adolescent population, where the research on NSSI is in its infancy.

Personality traits are a person’s fixed patterns of behavior and habitual ways of dealing with people in daily life [18]. Numerous studies have shown that personality traits are significantly correlated with suicide [19-22], and Erikson’s stages theory of personality development [23] proposes that each individual’s life can be divided into eight stages, and failure to resolve crises at each stage before adolescence increases the risk of suicide. Personality traits have greatly influence NSSI [24,25], and some studies have shown that the prevalence of NSSI in borderline personality disorder is as high as 80% [26], in addition, a cohort study by Grant F et al [27] showed that NSSI scores were higher in those suffering from borderline personality disorder than in the non-diseased group. However, the specific reasons for the interaction between personality traits and NSSI have been less explored for children and adolescents in the normal Chinese population. The Neuroticism Extraversion Openness Five-factor Inventory is a highly generalized and stable model that includes five dimensions of Neuroticism (N), Extraversion (E), Openness (O), Agreeableness (A), and Conscientiousness (C).

The main objective of this study was to explore the demographic factors that influence the severity of NSSI and to explore the role and possible causes of personality traits in the occurrence and severity of NSSI in five dimensions using the Neuroticism Extraversion Openness Five-factor Inventory.

## Methods

### Participants and procedure

The questionnaires were administered to a whole group of middle school students in three middle schools of PR China, written informed consent was obtained from participants and their guardians prior to the survey, and the questionnaires were organized by the class teachers of each class. The questionnaires were administered and collected after obtaining informed consent, and privacy was protected throughout the process. The collected questionnaires were then sorted, and those that were omitted, logically reviewed for errors, or did not meet the requirements were excluded, and data were entered and analyzed using SPSS 23.0 with a statistical test level of α=0.05. A total of 2900 questionnaires were distributed and 2376 valid questionnaires were returned (47.8% male, n=1136), with a validity rate of 81.9%, and the subjects’ ages ranged from 12 to 17 years (M age=13.66 years, SD=0.982).

### Measures

#### General situation questionnaire

A self-designed general information questionnaire was used to collect participants’ personal information, including gender (1 male, 2 female), age, grade, number of children in the family (1 single child, 2 multiple children) and upbringing area(1 urban, 2 rural).

#### Adolescents Self-Harm Scale

The Adolescents Self-Harm Scale developed by Ying Zheng et al [28] and revised by Yu Feng et al [29], has good reliability and validity and is widely used in China, the coefficient of internal consistency is 0.85. The questionnaire is a self-assessment questionnaire, and the scale lists 15 NSSI modalities (sharp cuts, burns, etc.) and one open-ended NSSI modality fill-in-the-blank, and investigates the number of NSSI occurrences in the past year, the degree of injury to the body, and the age at which the NSSI first occurred, the time of the most recent NSSI, the prompting cause or purpose, and the external stimulus event. The behavioral style, prompting cause or purpose, and external stimulus event of the NSSI can all be multiple choice, and subjects will answer as appropriate. There are 4 options for the number of occurrences of NSSI, which are 0, 1, 2-4, and ≥5, corresponding to 0, 1, 2, and 3 points. The degree of injury to the body has 5 options: none, mild, moderate, severe, and very severe, corresponding to 0, 1, 2, 3, and 4 points. In the degree of injury, “none” means no injury; “mild” means the injury can be healed without treatment, and the degree of injury does not affect daily life; “Moderate” means that the injury requires simple treatment by oneself or in a hospital, and the degree of injury has less impact on daily life; “severe” means that the injury requires more complicated treatment (such as stitches, etc.), and the degree of injury has more impact on daily life. “Very severe” means the need for hospitalization. The severity of NSSI was measured by the sum of the product of the frequency of each self-injury and the degree of injury, and the higher the score, the more severe the NSSI was.

#### Neuroticism Extraversion Openness Five-factor Inventory (NEO-FFI)

This questionnaire was initially developed by American scholars Costa and McCrae in 1978 (Neuroticism Extraversion Openness Personality Inventory, NEO-PI), and was later revised several times to form the Neuroticism Extraversion Openness Five-factor Inventory (NEO-FFI), which is widely used in various countries today, in 1992. The Chinese version of the questionnaire was revised by Professor Jianxin Zhang of the Chinese Academy of Sciences and is widely used in China to measure personality. The questionnaire consists of five dimensions: Neuroticism (N), Extraversion (E), Openness (O), Agreeableness (A), and Conscientiousness (C), with a total of 60 questions, all scored on a 5-point Likert scale, with “0” equals to “strongly disagree”, “1” equals to “disagree”, “2” equals “not sure”, “3” equals “agree” and “4” equals “strongly agree”. There are 27 reverse-scored questions in the questionnaire, and the sum of the scores of each question is the score of the dimension, and the higher the score, the more significant the personality characteristics of the respondent in that dimension. The questionnaire has good reliability and validity, the coefficient of internal consistency for the five dimensions of Neuroticism, Extraversion, Openness, Agreeableness, and Conscientiousness were 0.85, 0.80, 0.68, 0.75, and 0.83, respectively.

### Statistical methods

The sample was divided into groups with and without NSSI based on the presence or absence of NSSI in the past year, with a total of 881 individuals (56.6% female; n=499) with a mean age of 13.61 years (SD=1.017) in the group with NSSI and 1495 individuals (49.6% female; n=741) with a mean age of 13.69 in the group without NSSI (SD= 0.960). We described the general information and NSSI implementation methods of the two groups, compared the personality characteristics of the two groups using independent samples t-test, observed the correlation between personality characteristics and NSSI severity using Pearson correlation analysis, established scatter plots and fitted equations for personality characteristics and NSSI severity, and finally analyzed the severity of gender, age, grade, solitary status, upbringing, and personality characteristics with NSSI using stepwise regression analysis.

## Results

### General information

Of all subjects in this survey, 52.2% (n=1240) were female, 31.8%, 36.9%, and 31.3% (n=755, 877, and 744) were in the first, second, and third year of junior high school, respectively, and single-child households accounted for 13.1% (n=311) and urban households accounted for 12.0% (n=286). The study showed that the annual incidence of NSSI among junior high school students in China was 37.1% (n=881), which was similar to the results of previous studies (5.4%-44.3%). The mean age of occurrence was 13.61 years (SD=1.017), with females accounting for 56.6% (n=499) of the total number of detections. Among subjects who developed NSSI behaviors, the median self-injury level score was 4 (P25=1, P75=9).

### NSSI implementation

This survey showed that 32.6% (n=287) of the number of detections were self-injury using a single modality and 67.4% (n=594) were self-injury using a non-single means. Hair pulling was the most common mode of NSSI with an incidence of 51.0%. Cutting the skin with objects such as pocket knives and hitting harder objects such as walls with hands were also common NSSI modalities, with the number of people performing each modality accounting for 43.0% and 42.8% of the total number of detections (Table 1).

**Table 1.** NSSI method statistics.

| Implementation method | Number (n) | Percentage (%) |
| --- | --- | --- |
| Pulling hair | 449 | 51.0 |
| Cut skin with glass, knife, etc. | 379 | 43.0 |
| Hitting harder things such as walls or glass with your hands | 377 | 42.8 |
| Poking open wounds and preventing them from healing | 341 | 38.7 |
| Bleeding from scraping or rubbing the skin | 272 | 30.9 |
| Biting | 217 | 24.6 |
| Carve words or designs on your body (except for tattoos) | 166 | 18.8 |
| Scratching until there are bruises or bleeding | 143 | 16.2 |
| Stabbing something into skin or under fingernails | 119 | 13.5 |
| Lighting a fire in hand or touching a flame | 106 | 12.0 |
| Banging head against something until it bruises | 89 | 10.1 |
| Punching the body until it bruises | 58 | 6.6 |
| Bleeding from sticking body with needles, nails, etc. | 55 | 6.2 |
| Strangling with a string or other object to bruise | 48 | 5.4 |
| Burning/scalding skin with cigarette butts, etc. | 45 | 5.1 |
| Other | 77 | 8.7 |

Statistics on the way Chinese adolescents perform NSSI

### Comparison of personality traits between groups with and without NSSI

The scores of the group with NSSI in Neuroticism dimension were higher than those of the group without NSSI in the same dimension, while the scores in Extraversion, Openness, Agreeableness and Conscientiousness were lower than those of the group without NSSI in the same dimension (p < 0.01; Table 2).

**Table 2.**
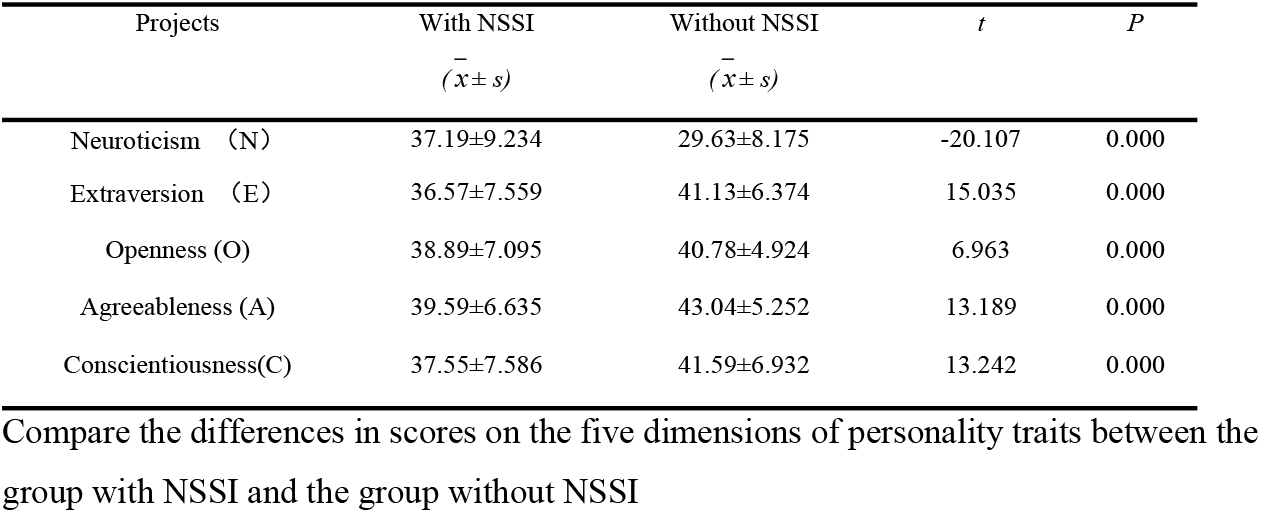
Comparing the differences in personality trait scores between the two groups.

| Projects | With NSSI<br>($\bar{x} \pm s$) | Without NSSI<br>($\bar{x} \pm s$) | *t* | *P* |
| --- | --- | --- | --- | --- |
| Neuroticism (N) | 37.19±9.234 | 29.63±8.175 | -20.107 | 0.000 |
| Extraversion (E) | 36.57±7.559 | 41.13±6.374 | 15.035 | 0.000 |
| Openness (O) | 38.89±7.095 | 40.78±4.924 | 6.963 | 0.000 |
| Agreeableness (A) | 39.59±6.635 | 43.04±5.252 | 13.189 | 0.000 |
| Conscientiousness(C) | 37.55±7.586 | 41.59±6.932 | 13.242 | 0.000 |

Compare the differences in scores on the five dimensions of personality traits between the group with NSSI and the group without NSSI

### Relationship between personality traits and NSSI severity

#### Correlation Analysis

Pearson correlation analysis of the dimensions of personality traits with NSSI severity in the sample with NSSI showed that Neuroticism was positively correlated with NSSI severity (r=0.423, p<0.001), Extraversion was negatively correlated with NSSI severity (r=-0.384, p<0.001), and Openness was positively correlated with NSSI severity (r=0.138, p <0.001), Agreeableness was negatively correlated with NSSI severity (r=-0.430, p < 0.001), and Conscientiousness was negatively correlated with NSSI severity (r=-0.338, p <0.001) (Fig 1).

**Fig 1.**
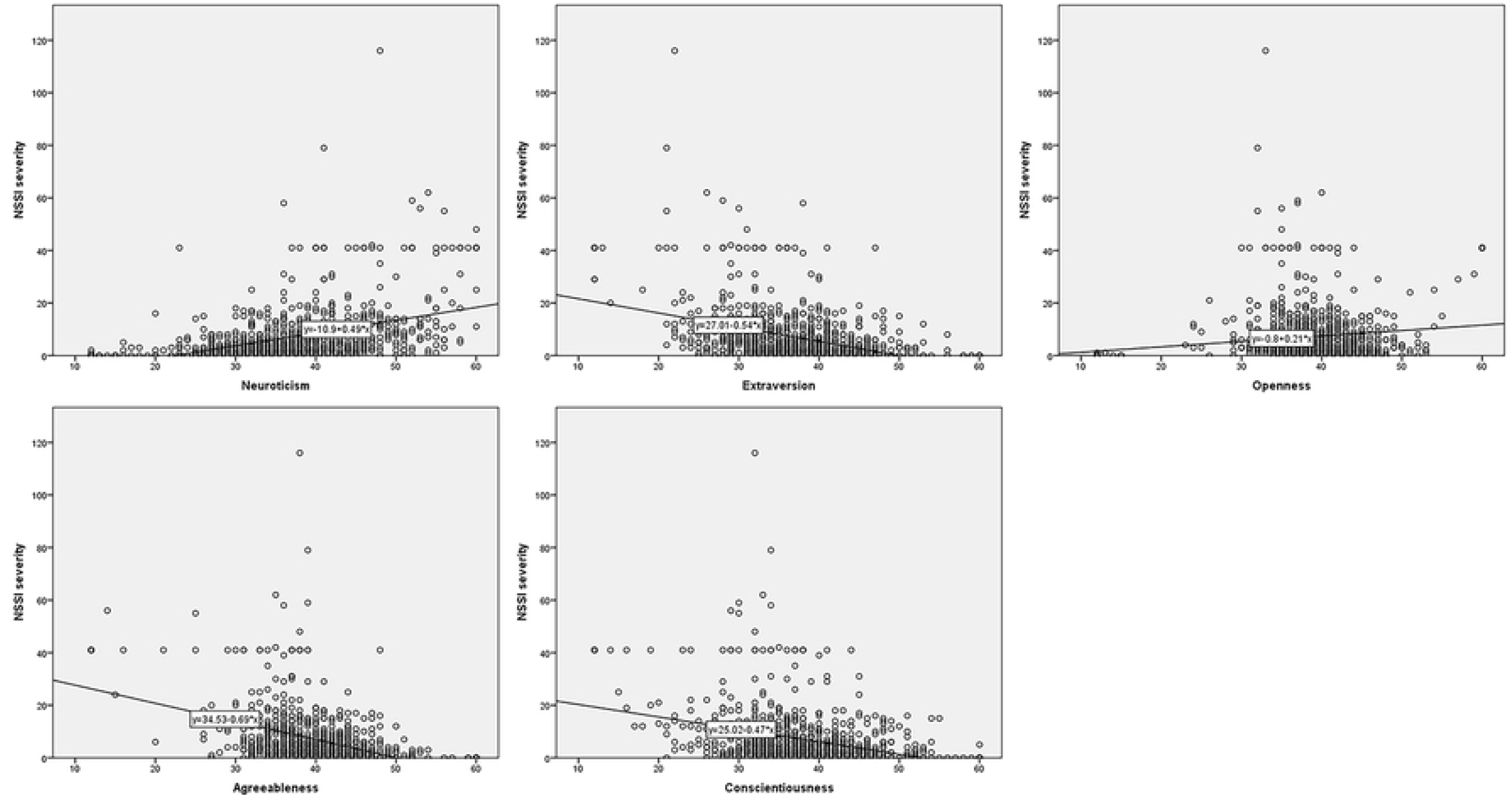
Correlation between dimensions of personality traits and NSSI severity. Using the dimensions of personality traits as horizontal coordinates and the severity of NSSI as vertical coordinates, scatter plots and fitting equations were constructed for the correlation between the two respectively.

#### Stepwise regression analysis

A linear regression equation was constructed using the “stepwise” method with age, gender, grade, number of children in the family, upbringing, and personality traits as independent variables and NSSI severity as dependent variables. The results showed that gender, Neuroticism, and Openness were positively associated with NSSI severity (p < 0.05), and grade, Extraversion, Agreeableness, and Conscientiousness were negatively associated with NSSI severity (p < 0.05; Table 3).

**Table 3.** Analysis of factors influencing the severity of NSSI.

| Dependent variable | Independent variable | Partial regression coefficient |  |  |  | Standardized partial regression coefficients | R<sup>2</sup> | R<sup>2</sup><sub>adj</sub> |
| --- | --- | --- | --- | --- | --- | --- | --- | --- |
| | | $\beta$ | SE | t | P | | | |
| NSSI severity | grade | -0.806 | 0.378 | -2.133 | 0.033 | -0.060 | 0.313 | 0.308 |
|  | gender | 1.364 | 0.620 | 2.201 | 0.028 | 0.064 | 0.309 | 0.305 |
|  | Neuroticism | 0.217 | 0.039 | 5.529 | < 0.01 | 0.188 | 0.260 | 0.258 |
|  | Extraversion | -0.210 | 0.048 | -4.366 | < 0.01 | -0.149 | 0.283 | 0.281 |
|  | Openness | 0.178 | 0.043 | 4.161 | < 0.01 | 0.119 | 0.294 | 0.291 |
|  | Agreeableness | -0.392 | 0.052 | -7.596 | < 0.01 | -0.245 | 0.185 | 0.184 |
|  | Conscientiousness | -0.182 | 0.046 | -3.976 | < 0.01 | -0.130 | 0.305 | 0.302 |

Multiple linear stepwise regression analysis was performed with NSSI severity as the
dependent variable and demographic characteristics information and personality traits as independent variables.

## Discussion

### General Information

Through our study, we found that the detection rate of NSSI among Chinese junior high school students in the past year was 37.1%. Previous surveys conducted on Chinese students aged 11-19 years [30] showed that the percentage of those who had at least one NSSI in the past year was 38.5%, similar to the results of the present study. In terms of demographic characteristics, the higher the grade level, the lower the severity of NSSI. The specific reasons for this have not been reported, and may be related to the fact that the higher the grade level, the higher the level of education received, the gradual improvement of self-values, the gradual increase in control and the more rational understanding of things occur. The severity of NSSI is higher among female gender than male, and previous studies on the incidence of NSSI have shown that it is higher in women than in men [31-33]. For the reasons of the phenomenon, the sexual control model of self-injury suggests that self-injurious behavior in adolescent females is related to first menstruation and adverse menstrual reactions, and that females counteract such adverse reactions by self-injury [34], while some scholars believe that the phenomenon arises because of the different ways in which men and women perceive the problem [35], with boys being more likely to deal with the problem faster through rational perception, ultimately leading to a higher detection rate in females than in males. As for the severity of NSSI, the specific reasons have not been reported in relevant studies and may be similar to the mechanism of the role of gender on the incidence of NSSI. In terms of self-injury mode, the proportion of those who adopted hair pulling as a mode of NSSI was 51.0%, which may be related to the high incidence of NSSI in the female population and the fact that this behavior is simpler and easier to take and less harmful, as the purpose and motivation of NSSI is mainly to release and manage negative emotions (92.7%), followed by control and influence others (60.7%) [36], not to cause serious consequences. Scratching the skin and hitting harder objects with the hand are likewise both relatively easy to think of and achieve self-injury.

### Personality

The present study showed that all five dimensions of personality traits were significantly correlated with the occurrence of NSSI, which is not identical to the findings of Mengrong Xu et al [37], probably due to the fact that the previous study population was adolescent patients with depressive disorders, whereas the present study was conducted on general middle school students. In addition, personality traits are significantly weakly correlated with the severity of NSSI, and the weak correlation may be due to the fact that personality traits, as a determinant of a certain trait of a person, are relatively stable and do not change significantly with time or events. The influence of personality traits on NSSI specifically in life is mainly reflected through the way individuals behave. Personality traits will have a certain influence on individuals’ behavior and tendency to do things, but this influence is relatively limited, and individuals’ behavior will be influenced by various external factors in addition to personality traits, and will not be completely governed by personality traits, thus leading to a weak correlation between the two.

### Neuroticism

The present study showed that high Neuroticism is a risk factor for the occurrence of NSSI, which is similar to the results of previous studies [24]. Neuroticism was significantly <u>and</u> positively associated with NSSI severity. The reason behind this phenomenon may be due to the fact that individuals who score higher on the Neuroticism dimension have poor emotional regulation, are emotionally sensitive and fragile, and are not good at effective communication. These individuals are impulsive, irritable, panic-stricken when in trouble or facing difficulties, deal with problems without consequences, and sometimes cope with negative emotions by venting, and this behavior tends to amplify the adverse effects of the problem, leading to greater stress and more problems. It has been shown that individuals with early personality traits such as impulsivity and aggression are more likely to develop NSSI [38], thus high Neuroticism groups are more likely to develop NSSI and the severity of NSSI tends to be more severe.

### Extraversion

High Extraversion was a protective factor for the occurrence of NSSI, and extraversion was significantly and negatively associated with NSSI severity. For the occurrence of NSSI, the present study was contrary to the results of previous studies in China [24] and the same as those in other countries [39-45]. The reason why the results of this study are contrary to those of previous Chinese domestic studies may be due to the fact that this phenomenon was assessed using the E subscale of the EPQ in previous studies, whereas the NEO-FFI used in this study differentiates the Extraversion and Openness dimensions, and classifies the dimensions of this phenomenon in a more detailed way. The reason for this phenomenon may be that individuals with high levels of Extraversion are lively and cheerful in their lives, have close contact with their surroundings and people, and are able to solve problems effectively through good communication or receive help from outside, which reduces the influence of negative emotions on them. These individuals are more likely to be understood and accepted by others in interpersonal interactions due to their Extraversion personality, and tend to be in the leader position of the group and more likely to gain self-satisfaction and self-existence in social interactions. Previous studies have shown [46,47] that individuals’ Extraversion levels predict their current and 10-year positive mood levels. Another study on the difference in attention to positive and negative signals by inside-out tendencies [48] showed that individuals with high introversion have a significant attentional bias towards negative emotional signals. Therefore, they are more sensitive to negative events in life and more likely to have negative emotions. Individuals with low levels of Extraversion are introverted and passive in their lives, not good at expressing their inner thoughts, and communicate less with others and friends, so individuals cannot vent their negative emotions effectively. At the same time, these individuals have relatively poor social skills and are often in a passive and vulnerable position in interpersonal communication, and are easily troubled by social problems.

### Openness

High Openness is a protective factor for the occurrence of NSSI, but Openness is significantly <u>and</u> positively associated with NSSI severity. This “divergence phenomenon” may be due to the fact that individuals with high Openness tend to have high Extraversion for the occurrence of NSSI, which leads to a more extroverted personality, broad interpersonal relationships, and openness to the unknown and the environment. Therefore, high Openness is a protective factor for the occurrence of NSSI. However, this openness also means that individuals like adventure and excitement, and are exploratory and inclusive. Therefore, for individuals who have developed NSSI, they are more likely to come into contact with other NSSI individuals in their lives, and are more likely to interact with each other, learn more about NSSI means, and even develop the “comparison phenomenon” of NSSI. Previous studies have revealed the phenomenon of social transmission [49], and the authors noted that groups with openness to external things are more likely to develop socially transmission behaviors, thus resulting in relatively higher levels of NSSI in individuals with high Openness.

### Agreeableness

High Agreeableness is a protective factor for the occurrence of NSSI, and Agreeableness is significantly and negatively associated with the severity of NSSI, which has not been previously reported. The reason may be due to the fact that individuals with high levels of Agreeableness are more compassionate, tolerant and trusting in interpersonal interactions. These individuals have a more broad-minded personality, are more tolerant of adversity and negative events, are more emotionally stable, and are less likely to have NSSI and less severe NSSI. This reminds us that cultivating qualities such as tolerance, empathy, trust, and stress tolerance in adolescents has positive implications for the prevention of behavioral problems.

### Conscientiousness

High Conscientiousness is a protective factor for the occurrence of NSSI, and Conscientiousness is significantly and negatively correlated with the incidence and severity of NSSI. Individuals with high levels of conscientiousness have greater control over their own behavior. They have a strong ability to organize their social activities and are more persistent in their actions. When encountering negative emotions, these individuals are able to analyze the problem rationally and solve it through reasonable and appropriate behaviors to alleviate their bad emotions. At the same time, such individuals are more self-disciplined, more able to achieve personal fulfillment in their social life, and therefore less likely to have NSSI and less severe NSSI. This reminds us whether the next step to investigate NSSI could be to measure neurotransmitters or electrical signals in the brain, starting from the behavioral inhibition function of the brain, to discover objective indicators that can predict NSSI.

## Conclusion

Despite some limitations, our study suggests that the annual incidence of NSSI remains high in the current sample. The severity of NSSI was relatively high in lower grades and females, and the number of self-injury using non-single means was higher, with hair pulling, skin scratching and whacking harder objects with the hands being the most common NSSI modalities. This provides a reference for future mental health interventions for middle school students and the prevention of NSSI behaviors. Personality characteristics have a significant correlation with the incidence of NSSI and the degree of self-injury, which requires that in future education of children and adolescents, different management and teaching methods for individuals with different personality characteristics can reduce the incidence of NSSI. For individuals who have developed NSSI, different measures targeted according to personality characteristics can effectively reduce the severity of NSSI, which is necessary for the healthy growth of junior high school students.

## Data Availability

All relevant data are within the attach files.

## Acknowledgments

We thank all the subjects who participated in this survey for their generous contributions to the study and all the classroom teachers of the participating classes for their active cooperation.

## Author Contributions

**Conceptualization:** Zhongliang Jiang, Qiang He, Xiaojing Cheng, Jie Chen, Geng Tian.

**Data curation:** Zhongliang Jiang, Zhiyi Wang, Miao Zhao, Long He.

**Formal analysis:** Zhongliang Jiang.

**Investigation:** Zhongliang Jiang.

**Supervision:** Jintong Liu, Xianbin Li, Muhammad Imran Haider.

**Writing – original draft:** Zhongliang Jiang.

**Writing – review & editing:** Zhongliang Jiang, Jintong Liu.

